# Pooled Saliva Specimens for SARS-CoV-2 Testing

**DOI:** 10.1101/2020.10.02.20204859

**Authors:** Bidisha Barat, Sanchita Das, Valeria De Giorgi, David K. Henderson, Stacy Kopka, Anna F. Lau, Tracey Miller, Theresa Moriarty, Tara N. Palmore, Shari Sawney, Chris Spalding, Patricia Tanjutco, Glenn Wortmann, Adrian M. Zelazny, Karen M. Frank

**Author notes:** Corresponding Author Karen M Frank, MD, PhD, Dept of Laboratory Medicine, Clinical Center, NIH, Room 2C306, Bldg 10, 10 Center Drive, Bethesda, MD 20892.

## Abstract

We evaluated saliva (SAL) specimens for SARS-CoV-2 RT-PCR testing by comparison of 459 prospectively paired nasopharyngeal (NP) or mid-turbinate (MT) swabs from 449 individuals with the aim of using saliva for asymptomatic screening. Samples were collected in a drive-through car line for symptomatic individuals (N=380) and in the emergency department (ED) (N=69). The percent positive and negative agreement of saliva compared to nasopharyngeal swab were 81.1% (95% CI: 65.8% – 90.5%) and 99.8% (95% CI: 98.7% – 100%), respectively. The sensitivity increased to 90.0% (95% CI: 74.4% – 96.5%) when considering only samples with moderate to high viral load (Cycle threshold (Ct) for the NP <=34). Pools of five saliva specimens were also evaluated on three platforms: bioMérieux NucliSENS easyMAG with ABI 7500Fast (CDC assay), Hologic Panther Fusion, and Roche COBAS 6800. The median loss of signal upon pooling was 2-4 Ct values across the platforms. The sensitivity of detecting a positive specimen in a pool compared with testing individually was 100%, 93%, and 95% for CDC 2019-nCoV Real-Time RT-PCR, Panther Fusion® SARS-CoV-2 assay, and cobas® SARS-CoV-2 test respectively, with decreased sample detection trending with lower viral load. We conclude that although pooled saliva testing, as collected in this study, is not quite as sensitive as NP/MT testing, saliva testing is adequate to detect individuals with higher viral loads in an asymptomatic screening program, does not require swabs or viral transport media for collection, and may help to improve voluntary screening compliance for those individuals averse to various forms of nasal collections.

## Introduction

A coronavirus outbreak (COVID-19) that was first reported in late December 2019 rapidly spread worldwide resulting in a pandemic. There are > 29 million SARS-CoV-2 infections and > 900,000 related deaths worldwide, with >6 million infections and >194,000 deaths in the United States (1). Screening, testing, and contact tracing are essential for patient management and to reduce further spread of disease. Diagnostic testing for SARS-CoV-2 has been challenging throughout the course of the pandemic for numerous reasons such as supply shortages. For symptomatic patients, a highly sensitive, specific, and reliably accurate assay is important, and the choice of specimen type can impact assay performance (2). The Centers for Disease Control and Prevention (CDC) currently lists the following upper respiratory specimen types as acceptable: nasopharyngeal swab, anterior nares, mid-turbinate, oropharyngeal (OP), and NP/nasal wash/aspirates, with the NP swab often considered the preferred method for diagnostic testing and the collection method to which other specimen types have been compared (3-5). However, there is inconvenience associated with NP and OP swab collection including patient discomfort (3, 6), some risk of exposure to healthcare personnel, the requirement for swabs, and the need for personal protective equipment (PPE). Alternative specimen sources, such as anterior nares, have been listed as an acceptable specimen type since early in the pandemic even though reported sensitivity is only about 86% (2). Saliva, however, which can be easily self-collected by patients and is non-invasive has not been studied adequately. The goals for SARS-CoV-2 testing in asymptomatic vs symptomatic individuals differ, with high participation rate and ease of collection being important considerations for screening an asymptomatic population. This is particularly relevant as there is an urgent desire to open schools and businesses and to promote economic recovery. At our institution, we have had frequent requests to offer saliva testing for employees who did not voluntarily agree to NP or MT collection because of a medical condition or personal aversion. We hope to engage these individuals in our voluntary screening program by providing a suitable alternative specimen type. When this study began, saliva was not an accepted specimen type, an Emergency Use Authorization was required by the Food and Drug Administration for testing saliva, and procurement of saliva collection devices with stabilizers was limiting. Previously published studies on saliva testing for COVID-19 vary from 71 to 100% in their reported percent positive agreement or sensitivity of saliva compared with NP (Table S1) (2-4, 6-19).

Importantly, the tested population, the saliva collection method, and the processing protocol have varied between the studies, making comparison of results challenging. The number of individuals tested in some studies was relatively low; therefore, performance of saliva warrants additional study to determine the robustness of saliva testing. Here, in a low-prevalence geographical region, we collected samples from a drive-through collection center for symptomatic or exposed employees and during ED visits to evaluate saliva for detection of SARS-CoV-2 infection, with a goal to add saliva as an option at our institution for asymptomatic employee screening. We also demonstrated that pooled saliva testing provides acceptable sensitivity on three separate platforms, two of which are high-throughput instruments.

## Methods

### Study Subjects

Subjects were enrolled at two sites. At the NIH, adult employees presenting to a drive-through testing center due to symptoms or exposure were invited to provide SAL at the time of the NP collection. Criteria for referral to the car line included symptoms consistent with potential COVID-19 after review by occupational medicine service or recent high-risk exposure to an individual known to be infected with SARS-CoV-2. After giving informed consent, participants were instructed to provide 3-5 mL of saliva using the drooling method into a sterile tube without any stabilizer or solution. Participants were asked to avoid coughing or clearing the throat, if possible, during the collection. Saliva was collected without restriction on timing or intake of food. Following the saliva collection, the NP swab was collected by a healthcare provider. Six participants who were known to be positive returned on subsequent dates and provided paired MT and SAL samples, avoiding the need for the potentially uncomfortable NP collection with an aim to improve study participation, for a total of seven MT specimens. At the Washington Hospital Center, subjects who presented to the emergency department with symptoms consistent with COVID-19 were invited to participate. The study was approved by the institutional review boards for both participating institutions.

### Specimen Collection and Processing

Saliva samples collected in sterile containers without additives were stored at 4°C until testing, and were tested within 36 hours of collection with residual volume from the samples being frozen at -70°C. NP samples were collected with flocked swabs (Puritan) into 3 mL of viral transport media (Corning) and were tested within 12 hours of collection. Saliva/NP/MT specimens (200 μL) were extracted using the NucliSENS easyMAG platform (bioMérieux, Marcy l’Etoile, France) resulting in 50 μL of eluate. All saliva samples were tested only at the NIH laboratory. If a saliva sample was thick and hard to pipet, it was treated with Mucolyse (ProLab Diagnostics, Richmond Hill, ON, Canada) 1:1 with heating at 35°C for 15 minutes. Following digestion, 400 μL was extracted by easyMAG for a 50 μL eluate. After testing of the specimens collected in the ED, the remaining NP samples were sent to the NIH laboratory for retesting on easyMAG/ABI 7500 platform, if specimen was available.

### SARS-CoV-2 Assay

Nucleic acid from individual specimens was extracted from 200 μL of Saliva/NP/MT specimens using the NucliSENS® easyMAG® platform (bioMérieux, Marcy l’Etoile, France) with an elution volume of 50 μL.

PCR was performed on the Applied Biosystems 7500 Fast Real-Time PCR System (Thermo Fisher Scientific, Waltham, MA) (20). The assay utilized primer/probe sets for nucleocapid protein, 2019-nCoV_N1 and 2019-nCoV_N2, and the human RNase P (RP) as an internal control to ensure that extraction and amplification was adequate as described. Cycle threshold (Ct) values were recorded for N1, N2 and RNAse P for each sample. Samples were considered positive for SARS-CoV-2 when both N1 and N2 targets were detected with Ct count <40. The positive signal for N1 or N2 alone was defined as an indeterminate result. The Panther Fusion® SARS-CoV-2 Assay is a real-time RT-PCR assay with detection of two conserved regions of the ORB1ab gene in the same fluorescence channel and was performed on the Panther Fusion (Hologic, Inc., San Diego, CA). The cobas® SARS-CoV-2 real-time RT-PCR test was performed on the cobas 6800 instrument (Roche Molecular Diagnostics, Pleasanton, CA). Amplification of SARS-CoV-2 target nucleic acid is achieved by the use of a two-target RT-PCR, one from the SARS-CoV-2 specific ORF1 a/b non-structural region (target 1) and one from a conserved region of the envelope E-gene common to all SARS-like coronaviruses (pan-Sarbecoviruses) (target 2). The pan-Sarbecovirus detection sets will also detect the SARS-CoV-2 virus. Specimens collected in the ED were tested on one of the platforms at the MedStar Washington Hospital Center Laboratory: BioGX SARS-CoV-2 Reagents for BD MAX(tm) System (Franklin Lakes, NJ, USA), Xpert^®^ Xpress SARS-CoV-2, Cepheid’s GeneXpert^®^ Systems (Sunnyvale, CA, USA), DiaSorin Molecular Simplexa(tm) COVID-19 Direct real-time RT-PCR, LIAISON® MDX instrument (Stillwater, MN, USA) or sent to a reference laboratory that uses the QuantStudio (Thermo Fisher, Waltham, MA).

### Pooling Saliva

Equal volumes of saliva from five subjects were pooled into a single tube. Proteinase K, 20 mg/mL (Invitrogen by Thermo Fisher Scientific, Waltham, MA) was added at a ratio of 12.5 μL per 100 μL volume, followed by vortexing, heating for 5 minutes at 95°C, and brief centrifugation. The following volumes of supernatant were loaded onto three different platforms: 400 μL onto NucliSENS easyMAG (bioMérieux, Marcy l’Etoile, France), 500 μL onto the Panther Fusion (Hologic, Inc., San Diego, CA), and 600 μL onto the COBAS 6800 (Roche, Pleasanton, CA). Individual samples that were thick were excluded from pooling and run as individual samples only, so none of the samples in the pool were treated with mucolyse prior to pooling.

### Statistical Methods

Wilcoxon signed-rank test was used to compare the cycle threshold (Ct) values. The 95% confidence intervals were calculated using the hybrid Wilson/Brown method. The correlation of Ct values between NP/MT and saliva was assessed using Pearson correlation coefficient and represented graphically with linear regression. A two-tailed T test with p <0.05 was considered statistically significant. The negative RT-PCR of the target gene was set at the Ct value of 40 for the statistical analysis. The NP swab test result was used as the reference method for the assessment of test agreement. For analysis of age range, 448 subjects of 449 were included because one subject’s age was not available. All statistical analyses were performed using GraphPad Prism version 8 (GraphPad Software, San Diego, CA). Only Ct values derived from testing on a single platform at NIH were included in the statistical analysis, with the only exception being the one calculation of percent positive agreement for the subset of higher viral load specimens (Ct <=34) for which the results from all platforms were considered, if the NP specimen was not also tested at NIH.

## Results

This study includes a total of 918 specimens (459 pairs) collected from 449 individuals between July, 13, 2020 and September 18, 2020. Of the total, 390 paired sets were collected from the NIH drive-through testing center and 69 were collected from the MedStar Washington Hospital Center ED. Participants in the drive-through testing center were symptomatic or had a recent high-risk COVID-19 exposure, and all patients in the ED had symptoms suggestive of possible COVID-19. The median age of participants was 42 (range 21 – 88 years), with 59% female, 41% male (Table S2). Of the 459 saliva samples, 75 were thick (57/390 (15%) from the drive-through and 18/69 (26%) from the ED) and were treated initially with mucolyse prior to individual testing. A total of 18 failed the initial extraction (13/390 (3%) from drive-through and 2/69 (3%) from the ED) and testing was repeated (Tables S3, S4). The percent positive and negative agreement of saliva compared to reference collection of NP/MT swab (440 NP and 7 MT) were 81.1% (95% CI: 65.8 % - 90.5%) and 99.8% (95% CI: 98.7% - 100%) respectively (Table 1). When considering samples with moderate to high viral load only, excluding the lower viral load specimens (defined as Ct of NP/MT <=34), the percent positive agreement increased to 90.0% (95% CI: 74.4% – 96.5%). See Table S5.

**Table 1.** SARS-CoV-2 Real-time RT PCR results for paired NP/MT and saliva

|  | NP<br>Positive/Indeterminate | NP Negative | Total |
| --- | --- | --- | --- |
| Saliva Positive/<br>Indeterminate | 30 | 1 | 31 |
| Saliva Negative | 7 | 421 | 428 |
| Total | 37 | 422 | 459 |

A comparison of the Ct of N1 for NP/MT and SAL for all samples tested on the NIH platform showed a higher viral load in the NP/MT samples compared to the SAL samples with median Ct of 26 for NP/MT compared to 31 for saliva (Figure 1A, 1B). Similar results were obtained upon comparison of N2 results for NP and SAL (Figure S1). There was a moderately good correlation of NP/MT Ct values with matched saliva (Figure S2). There was very good correlation for the N1 and N2 Ct values for both NP/MT and SAL (Figure S3A, S3B). Our analysis of the Ct values for the control RP gene indicates that the samples of different specimen types were adequate and the difference in Ct values of saliva vs NP/MT are not due to differences in human material obtained during the collection as saliva had slightly lower median Ct, meaning slightly stronger RP signal even though the SARS-CoV-2 signal is slightly less for saliva (Figure 1C).

**Figure 1 (A-C).**
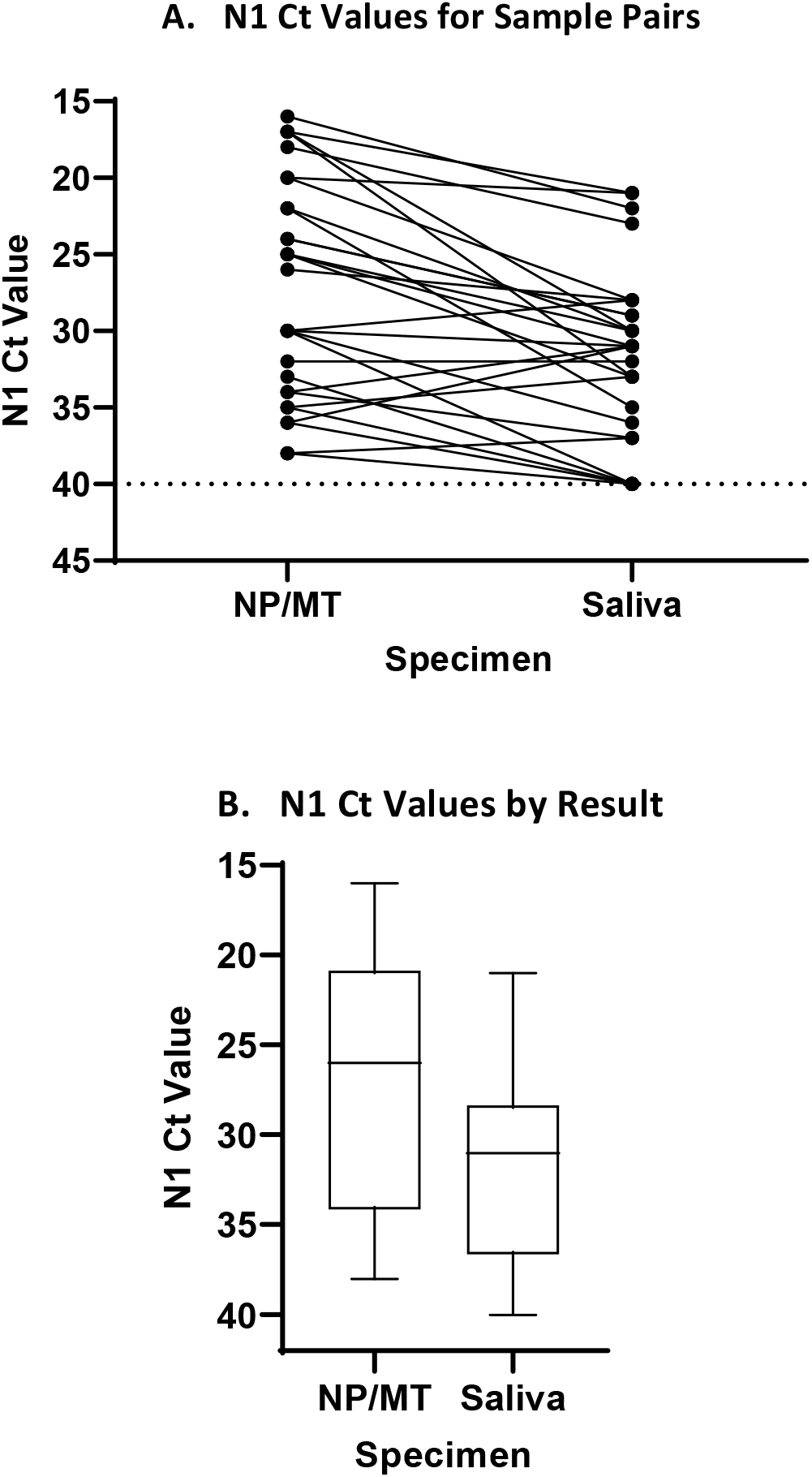
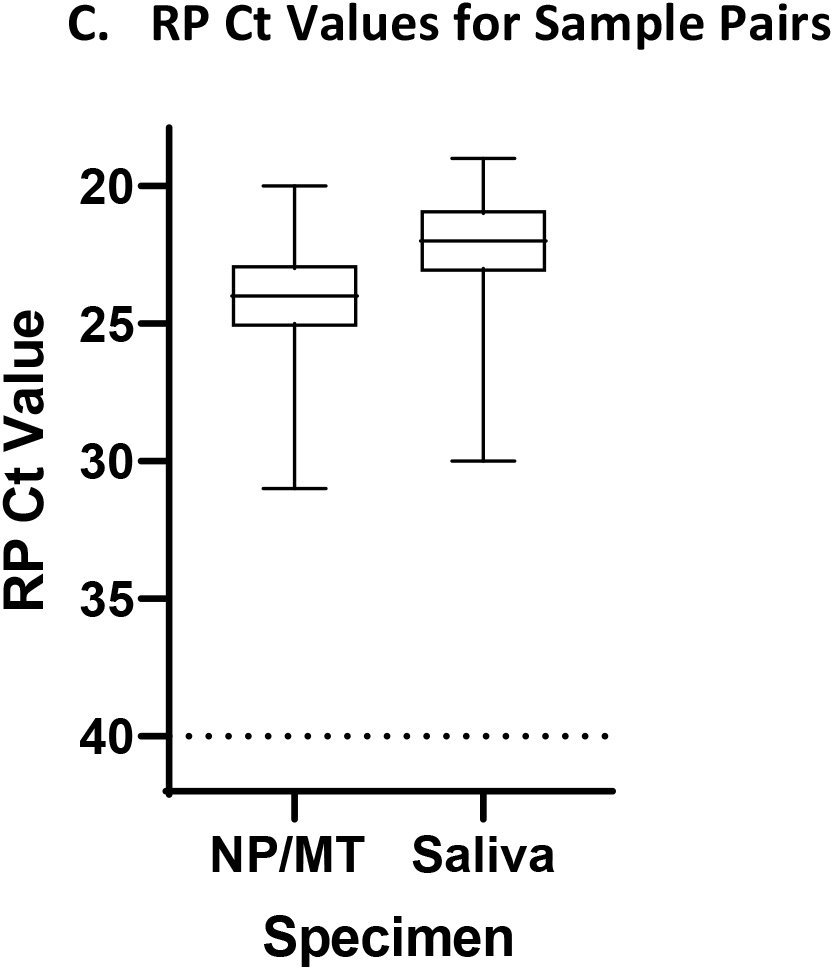
Comparison of Cycle Threshold (Ct) values of N1 for NP versus SAL specimens. A. N1 Ct values for paired NP/MT and SAL samples (29 pairs). Pairs are connected by a line. The N1 Ct was set to 40 for samples for which N1 was not detected, indicating negative for SARS-CoV-2 RNA. Horizontal dashed line is at Ct=40, the assay cut off. P-value < 0.001 calculated by Wilcoxon matched-pair signed rank test. B. A lower median viral load was seen for SAL specimens compared with the median Ct for NP/MT samples. Median and interquartile range are 26, (21-34) for NP/MT and 31, (29-37) for SAL respectively. P-value <0.001. C. RP Ct values for NP/MT and SAL specimens (424 pairs). Median and interquartile range are 24, (23-25) for NP/MT and 22 (21-23) for saliva respectively. Horizontal dashed line is at Ct=40, the assay cut off. P value < 0.001 calculated by Wilcoxon matched-pairs signed rank test.

To evaluate the pooling approach to testing, equal volumes of saliva were combined into a single tube, excluding samples too thick to pipet well, followed by treatment with proteinase K (21, 22). Three different platforms were tested to increase our options for automated workflow for screening, the CDC assay on the bioMérieux NucliSENS easyMAG/ABI 7500Fast platform, the Hologic Panther Fusion, and Roche COBAS 6800. For pooled testing on any platform, the results of the pool were compared to the individual saliva samples tested on the easyMAG/ABI 7500 platform, as that was our gold standard in the lab for individual saliva testing. For a pooled sample, the average loss of signal was 2-4 Ct values when compared with the individual sample for each platform (Figure 2A-C, Table S6). The sensitivity of detecting a positive specimen in a pool compared with testing individually was 100%, 93%, and 95% for easyMAG/ABI 7500, Hologic Panther Fusion, and Roche COBAS 6800 respectively, with decreased detection of samples with lower viral load as expected. The correlation of Ct values for individual samples versus pooled samples was slightly better for the CDC assay than for the Panther or COBAS assays (Figure S4 A-C). It is possible that future optimization of the processing steps for the automated platforms may lead to improved sensitivity.

**Figure 2 (A-C).**
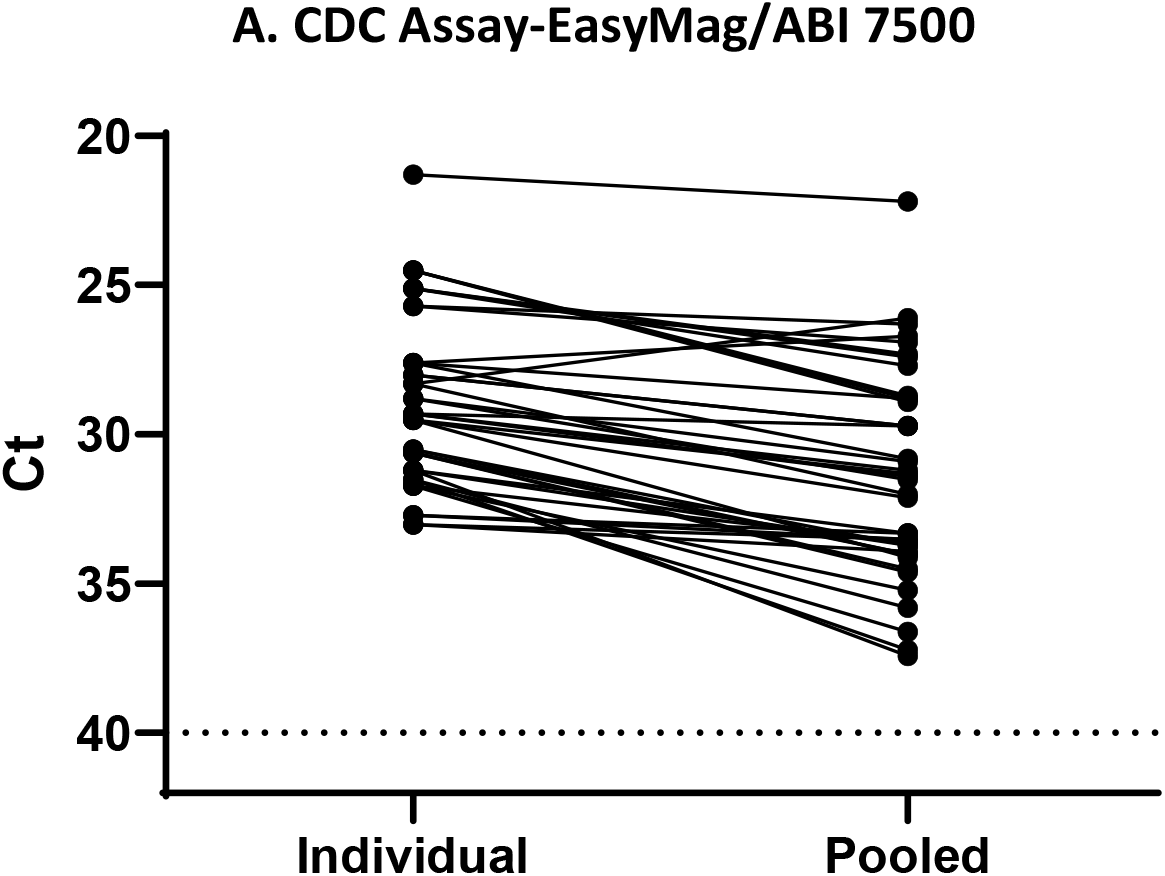
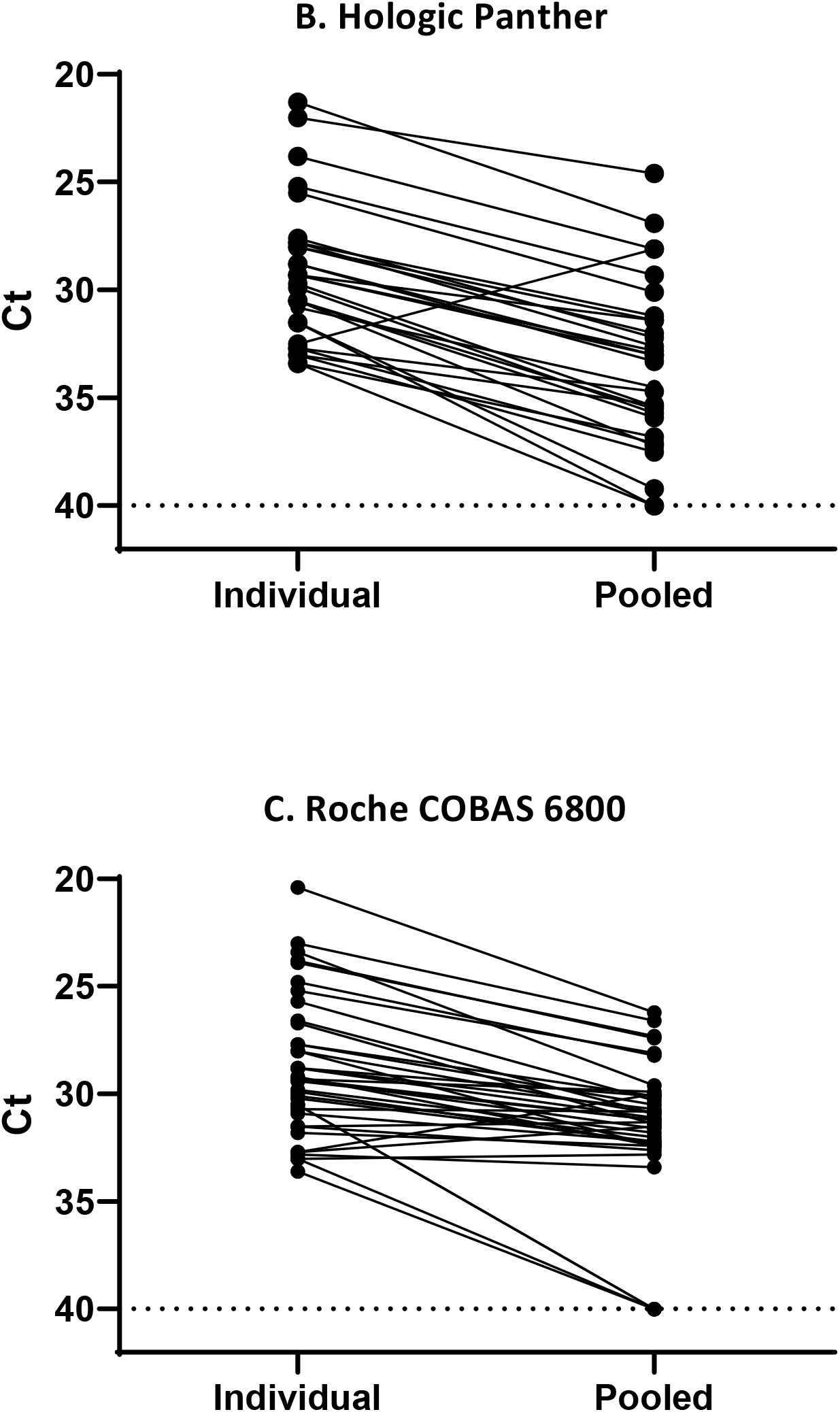
Comparison of Cycle Threshold (Ct) values for individual and pooled saliva specimens on different testing platforms. A. Ct values for paired individual and pooled samples (easyMAG/ABI 7500) for 41 pairs. B. Ct values for paired individual (easyMAG/ABI 7500) and pooled samples (Hologic Panther) for 30 pairs. C. Ct values for paired individual (easyMAG/ABI 7500) and pooled samples (Roche COBAS 6800) for 39 pairs. For A-C, pairs are connected by a line. Horizontal dashed line is at Ct=40, the assay cut off. P-value < 0.001 calculated by Wilcoxon matched-pair signed rank test. For C,D, the pooled Ct was set to 40 for samples in which N1 was not detected including those negative for SARS-CoV-2 RNA.

## Discussion

With the unprecedented number of deaths worldwide due to a coronavirus infection, screening, testing, and contact tracing for SARS-CoV-2 are essential. Developing new diagnostic measures for detection of COVID-19 is of critical importance to meet the global public health needs of COVID-19 testing. Because saliva can be self-collected, specimen collection can be simplified whereby the number of health care professionals in PPE in special collection centers can be reduced (4, 6, 23). Beginning May 2020, the NIH instituted a program to test asymptomatic employees weekly, but voluntary participation rate was far lower than desired. Some individuals found the NP or MT collection too uncomfortable for routine testing on a weekly basis. The goal of this study was to evaluate and add saliva as an alternative testing option for NIH employee asymptomatic screening only; not to replace our existing test algorithm for symptomatic patients. However, given the low rate of infections identified through our asymptomatic testing program (0.1% positivity rate), we enrolled symptomatic and high-risk exposed individuals through our drive-through collection site (5% positivity rate) and from a local ED (23% positivity in our study set). During the course of the COVID-19 pandemic, individual laboratories have been required to validate many different platforms due to supply shortages, multiple collection devices, and various specimen types. Although there are a number of published studies comparing specimen types, each study has a limited number of subjects and there are variations in collection methods, participant characteristics, and testing platforms. In order to be approved to conduct saliva testing, based on regulatory guidelines at the time, we were required to compare paired NP and saliva collections from the same individuals, not only to validate saliva as an acceptable specimen type on our instrument.

The range of reported sensitivity or percent positive agreement of the saliva collection method, most often compared to NP swab, varies widely from 71 to 100% and is too broad to make a specific guideline without further refinement of the analysis (2-4, 6-19). While our study and others show the acceptability of testing saliva, important variables need to be considered when reviewing various reported conclusions. These include severity of disease (asymptomatic to severe disease in hospitalized patients), method of collection (collection upon waking before any food or water intake, versus forced cough collected later in the day, versus drooling technique with no restriction on food/water intake at a random time later in the day), the gold standard or reference method for comparison in each study (NP versus NP/OP, versus MT), healthcare provider collected versus self-collected NP, addition of stabilizing agent, processing steps, RNA extraction process, and testing platform. Each of the studies alone is limited by which group of individuals was tested, the time and method of collection, and processing methods (4, 19). Some studies were limited by the inability of individuals to elicit a cough when requested (14), and there is a need to consider potential preanalytical errors caused by home-collected samples. It is possible that viral RNA extraction as well as RT-PCR efficiency might differ with the use of different preservation solutions based on their ability to protect viral RNA from degradation as well as their extraction chemistry (24).

Some studies have shown a lower viral load in saliva (13, 16, 17), but other studies showed similar viral loads between specimens or better viral loads in saliva (6). Studies have reported that higher viral loads were seen in patients with more severe disease (6, 7). In our study, the Ct values were on average higher in saliva (indicating a lower viral load) compared with NP. Comparison of first morning saliva versus a randomly timed collection was not an option for our study, given the consenting workflow. Saliva samples may be less optimal when not a first morning collection, for asymptomatic individuals, for those without food/water restriction, and for those later in the course of disease. Importantly, the range of viral load in the specimens in a small study can greatly affect the final calculated percent positive agreement because the specimens with higher viral loads are more likely to be detected by both NP/MT and SAL; therefore, studies with a higher median viral load across most specimens will show higher percent positive agreement than a study with a lower median viral load. The percent positive agreement on our study changed from 81% to 90% when only moderate to high viral load samples were included in the analysis. A meta-analysis that accounts for collection methods, patient population, and processing methods will lead to a more comprehensive understanding of the usefulness of SARS-CoV-2 saliva testing.

In order to provide high volume screening using saliva, there was a decision at our institution made to pool the samples. We had previously demonstrated that pooling of ten NP samples resulted in only a slight drop in sensitivity (losing an average of 3 Ct values) (25). For saliva, we chose to pool only five saliva samples because the saliva specimen as collected already resulted in a lower sensitivity. When pooling was applied, sensitivity was 100%, 93%, and 95% for the easyMAG/ABI 7500, Panther Fusion, and COBAS 6800, respectively. To date, only a few studies have evaluated the pooling of saliva (26, 27). Pooling conserves reagents and allows for higher throughput. The difference in Ct values between individual saliva samples and pooled saliva samples was 2-4 in our study. When combined with the lower rate of detection of infected individuals using saliva in our study, one might conclude that the use of pooled saliva on an automated platform, albeit with a slightly lower sensitivity, might be acceptable to promote compliance for screening.

The limitations for our study included the low number of positive participants, testing of symptomatic patients to determine an approach for screening the asymptomatic population, and the combined use of two collection sites (drive-through center and ED). The positive specimens include seven MT of the total 38 positives, to increase likelihood of participation in the study. All positive NP samples from the ED did not have a Ct value from the easyMAG/ABI 7500 platform, as not all samples were available for repeat testing. For this reason, only data from the easyMAG/ABI 7500 platform are included in the figures that compare Ct ranges.

A challenge for all centers offering saliva testing is that some individuals may have difficulty producing adequate saliva for the test. Saliva is also a more challenging specimen for the laboratory staff to handle and requires judgement about thickness to ensure the correct volume is pipetted, with a chance of an under-pipetted sample, due to viscosity or bubbles, leading to a false-negative result, as well as increased likelihood of extraction failure. Initially, mucolyse was added to individual thick saliva specimens prior to extraction, but data obtained during our pooling validation showed that proteinase K digestion for individual thick samples prior to extraction was just as effective. Therefore, thick specimens and pooled specimens follow the same processing procedure.

When evaluating the effectiveness of saliva collection, it is important to define which individuals are to be captured by the testing. Is the goal to detect anyone who has an infection with the virus or to detect those more likely to be infectious, reported to be Ct <35 in several studies (28-30), with other studies reporting as low as <24 (31). When comparing across published studies, the agreement between reports might increase if considering only samples with higher viral load. For these cases, the consensus appears to be that saliva is an acceptable and convenient method of testing. We conclude that saliva testing would detect employees who were most likely to be infectious to others and that saliva would be an adequate screening approach, although we encourage employees to opt for mid-turbinate collection, if they are willing, as it appears to be a more sensitive approach. Saliva testing is not used for individual patient diagnosis at our institution.

## Data Availability

All data for this study are included in supplemental tubes S3-S6.

## Acknowledgments

We thank the NIH car line team for assistance with sample collection, the clinical laboratory technologists in the Departments of Laboratory Medicine and Transfusion Medicine for performing assays, and the departmental support staff for managing paperwork related to the IRB protocol. We thank Masashi Waga and the MedStar Washington Hospital Center laboratory staff for providing lab data and samples after routine testing, and Tricia Coffey, Karen Kaczorowski, and Amanda Grove for assistance with patient registration. We would like to thank Larry Tabak, Janice Lee, Blake Warner, and Pavel Khil for helpful discussions.

KMF, DKH, SK, T Miller, TNP, study conception and design; KMF, BB, SD, AFL, TNP, AMZ, T Moriarty, SS, PT, GW collection of samples; KMF, SD, AFL, AMZ, VD, supervision of assays; KMF, BB, SD, CS, collection of data, performance of data analysis, and interpretation of data; KMF, BB, manuscript writing; AFL, TNP, SD, SK, VD, AMZ, manuscript editing. All authors reviewed and approved the manuscript.

We declare that we have no conflicts of interest. This work was supported in part by the Intramural Research Program of the National Institutes of Health Clinical Center. This project has been funded in part with federal funds from the National Cancer Institute, National Institutes of Health, under Contract No. 75N910D00024, Task Order No., Task Order No. 75N91019F00130. The content of this publication does not necessarily reflect the views or policies of the Department of Health and Human Services, nor does mention of trade names, commercial products, or organizations imply endorsement by the U.S. Government.

