## Supplemental Data for "Pooled Saliva Specimens for SARS-CoV-2 Testing"

**Barat et al.  Supplementary Data**

**Table S1: Summary of studies evaluating saliva as a specimen for SARS-CoV-2 testing**

| Reference, First Author | Characteristics | Collection Method | Total Subjects | Total Positive | Sensitivity/ PPA (%) SAL/ NP | Specificity / NPA (%) SAL/NP | Testing Platform and Notes |
|---|---|---|---|---|---|---|---|
| Jamal (1) | All known positive | Saliva<br><br>NP swab | 91 | 72 | 72 SAL<br><br>89 NP | - | Allplex™ 2019-nCoV Assay(100T) (Seegene Inc, Seoul, Korea)<br><br>Saliva + PBS |
| Nagura-Ikeda (2) | 15 asymptomatic,<br><br>88 symptomatic<br><br>All known positive | Saliva<br><br>OP/NP for diagnosis | 103 | 103 | SAL<br><br>82 LDT<br><br>81 COBAS<br><br>71 RT-LAMP | - | RT-qPCR LDT, cobas SARS-CoV-2 high-throughput system, RT-LAMP[1]<br><br>Saliva + PBS |
| Wyllie (3) | In-patient<br><br>Asymptomatic health-care workers | Saliva upon waking prior to food/water intake<br><br>NP swab<br><br>Self- collected NP for asymptomatic[2] | 70<br><br>495 | 70<br><br>9 | 81 SAL<br>71 NP<br><br>Only 9 positive samples for comparison | - | CDC Assay<br><br>Viral load higher in saliva in patients |

| | | | | | | | |
|---|---|---|---|---|---|---|---|
| Migueres (4) | 17 Asymptomatic<br><br>27 Symptomatic<br><br>9 Hospitalized | Saliva<br><br>NP swab | 123 | 44 | 88 SAL asymptomatic<br><br>95 SAL symptomatic<br><br>93 NP | - | Panther Fusion™ module (Hologic) |
| Skolimowska (5) | Symptomatic healthcare workers<br><br>5 pediatric patients (<18 years) | Saliva<br><br>Combined OP/NP swab comparison | 132 | 18 | 83 SAL | 99 SAL | Roche, AusDiagnostics, ThermoFisher, Abbott<br><br>cobas ® PCR medium |
| Pasomsub (6) | Symptomatic | Saliva<br><br>NP and throat swab | 200 | 19 | 84 SAL | 99 SAL | SARS-CoV-2 Nucleic Acid Diagnostic Kit (Sansure, Changsha, China)<br><br>CFX96 Real-Time Detection System (Bio-Rad) |
| Williams (7) | Screening clinic | Saliva<br><br>NP swab | 522 | 39 | 85 SAL | 98 SAL | Coronavirus Typing (8-well) assay, AusDiagnostics)<br><br>Higher viral load in NP |
| Zhu (8) | | Saliva<br><br>Respiratory tract sample (NP /OP swab) | 944 | 442 | 86 SAL | 97 SAL | |

| | | | | | | | |
|---|---|---|---|---|---|---|---|
| Landry (9) | Symptomatic out-patient | Saliva<br><br>NP swab | 124 | 35 | 86 SAL<br><br>94 NP | 98 SAL | CDC Assay<br><br>1/3 saliva samples were thick |
| Rao (10) | Asymptomatic adult male, 8-10 day after tested positive<br>SAL from deep in throat upon waking prior to food/water intake | Saliva<br><br>NP swab | 217 | 160 | 93 SAL<br><br>53 NP | - | One-step RT-PCR , Real-Q 2019 nCoV detection kit (Biosewoom, Inc, South Korea)<br><br>Higher viral load in saliva |
| Hanson (11) | Drive-thru test center | Saliva<br><br>NP swab | 354 | 81 | 94 SAL | 98 SAL | Hologic Aptima SARS-CoV-2 transcription mediated amplification (TMA) assay<br><br>Saliva + Universal transport media |
| McCormick-Baw (12) | Emergency department | Saliva<br><br>NP swab | 156 | 50 | 96 SAL | 99 SAL | Cepheid Xpert Xpress SARS27<br>CoV-2 PCR test |
| Procop (13) | Screening Center | Elicited cough and saliva<br><br>NP swab | 216 | 38 | 100 SAL PPA | 99 SAL NPA | Aptima™ SARS-CoV-2 transcription mediated amplification assay (Panther™ System, Hologic) |

| | | | | | | | |
|---|---|---|---|---|---|---|---|
| Azzi (14) | Confirmed COVID-19 patients with severe or very severe disease | Saliva (drooling technique) Or Endotracheal tube<br><br>NP swab | 25 | 25 | 100 SAL | - | Abi Prism 7000 sequence detection system (Applied Biosystems)<br><br>QIAmp Viral RNA mini kit (Qiagen)<br><br>Luna Universal qPCR Master Mix (New England BioLab) |
| Iwasaki (15) | Symptomatic | Saliva<br><br>NP swab | 76 | 9 | 97 Overall percent agreement | - | One-Step Real-Time RTPCR Master Mixes and tepOnePlus Real Time PCR System (Thermo Fisher Scientific) |
| Uwamino (16) | 32 hospitalized patients<br><br>115 symptomatic staff | Saliva<br><br>NP swab | 147 | 32 | 87 Overall concordance (SAL & NP)<br><br>96 overall concordance within first 10 days | - | LightCycler96 (Roche, Basel, Switzerland)<br><br>2019 Novel Coronavirus Detection Kit (Shimaszu, Kyoto, Japan) |
| Byrne (17) | Emergency Department<br><br>In-patient symptomatic | Saliva<br><br>Nasal and throat swab | 110 | 14 | 86 SAL | - | QIAmp Viral RNA Mini Kit (QIAGEN)<br>Genesig Real-time Coronavirus COVID-19 PCR<br>Similar viral load |

[1]RT-LAMP : Reverse transcription-loop-mediated isothermal amplification
[2]All NP/OP healthcare collected except when noted

**Table S2: Study Participant Characteristics**

| | ED | Drive- Through | Total |
|---|---|---|---|
| Number of Participants | 69 | 380 | 449 |
| Gender, Male N (%) | 23 (33%) | 159 (42%) | 182 (41%) |
| Age, Range (median) | 21-88 (46) | 21-75 (42) | 21-88 (42) |

Figure S1.

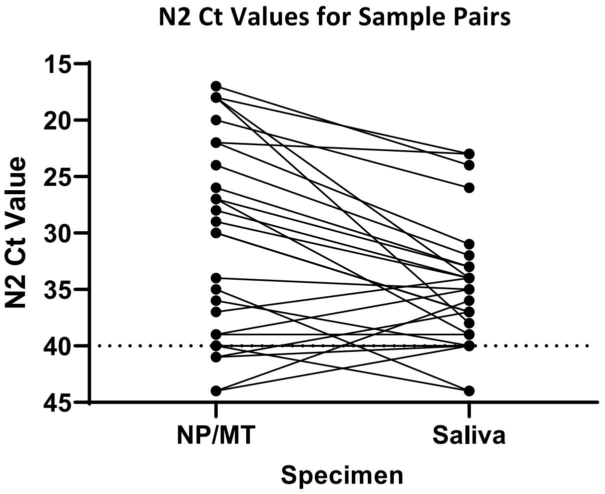


**Figure S1. N2 Cycle threshold (Ct) values for paired NP/MT and SAL samples (29 pairs).** Pairs are connected by a line. The N2 Ct was set to 40 for samples in which N2 was not detected, indicating a negative result for SARS-CoV-2 RNA. Horizontal dashed line is at Ct=40, the assay cut off. P-value <0.05 calculated by Wilcoxon matched- pair signed rank test.

Figure S2.

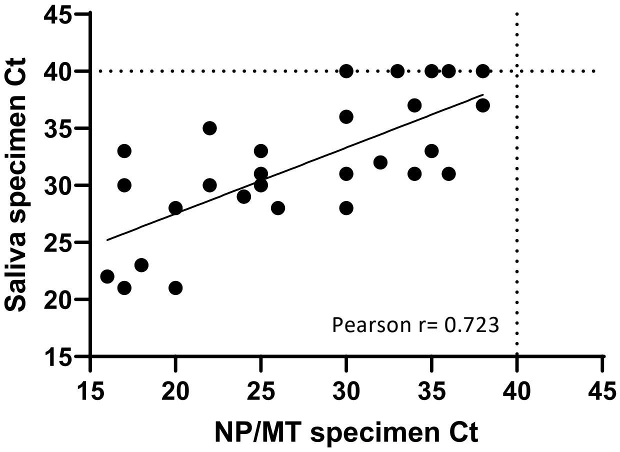


**Figure S2. Comparison of Cycle Threshold (Ct) values for N1 for NP versus SAL specimens (29 pairs).** Analysis demonstrates moderately good correlation between NP Ct values and Ct values for matched SAL samples. R and p values determined by Pearson correlation and graphically represented by linear regression. Dotted line indicates the limit of detection. P-value <0.001.

Figure S3 (A-B)

**A. NP/MT Samples**

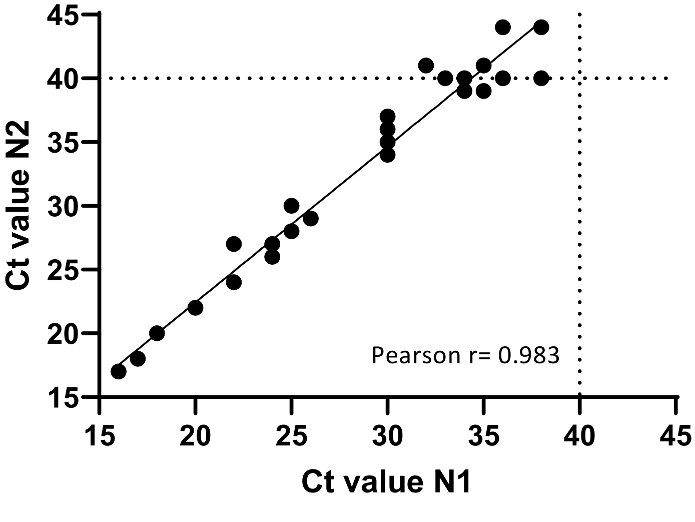

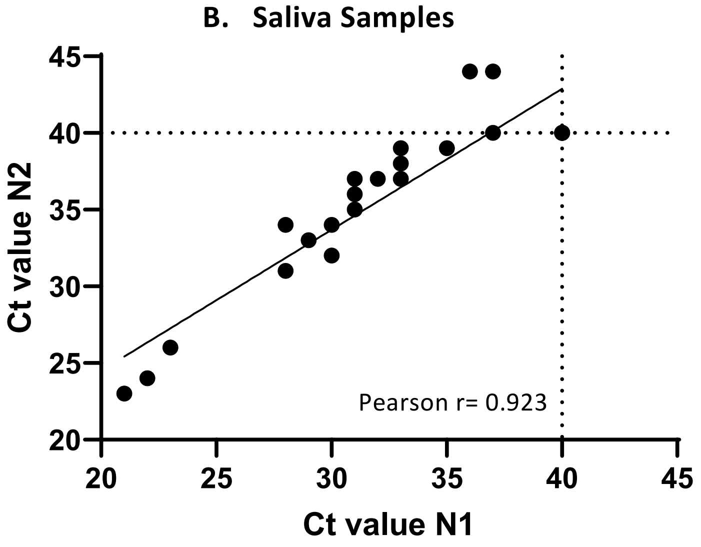


**Figure S3 (A-B). Concordance between SARS-CoV-2 detection in 29 samples of each type using CDC N1 primer and probe sets.** R and p values determined by Pearson correlation and graphically represented by linear regression. Dotted line indicates the limit of detection. P-value <0.001

Figure S4 (A-C)

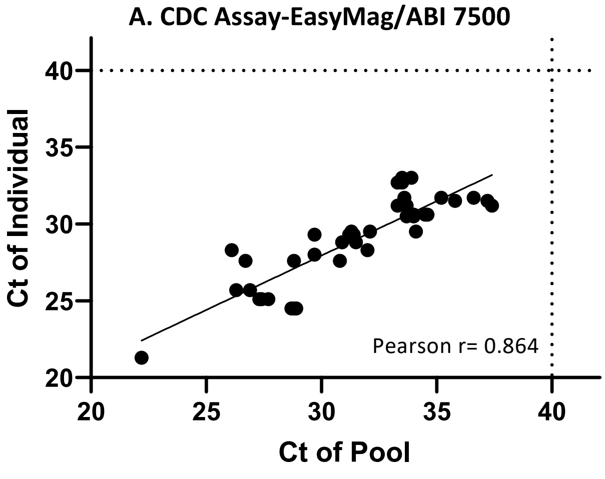

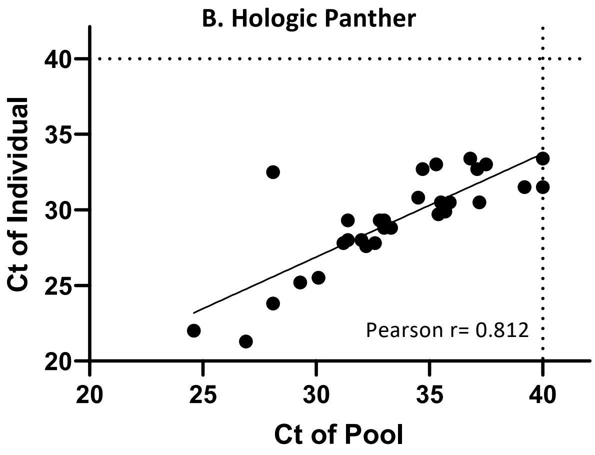


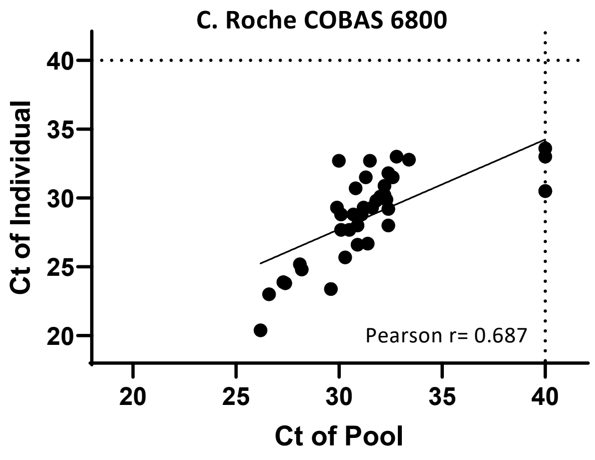


**Figure S4 (A-C). Relationship between Ct values of individual and pooled SAL samples.** R and p values determined by Pearson correlation and graphically represented by linear regression. Dotted line indicates the limit of detection. P-value<0.001
